# Pilot application of the BPSConnect model: bio-psycho-social correlation in children with ASD

**DOI:** 10.1101/2025.09.24.25336461

**Authors:** Rebeca Linares, Sindia Linares Castellanos, Norma Mas Vallespin, Maialen Lancho Azurmendi, Anyela Tobar Guerrero, Marina Linares Crespo

**Affiliations:** e-TherapyKids, Barcelona, Spain; Independent Clinical Nutritionist; e-TherapyKids Clinic

**Keywords:** Autism spectrum disorder, bio-psycho-social model, biomarkers, epigenetics, parental stress, BPSConnect, pilot study

## Abstract

**Objective:** To evaluate the feasibility and preliminary clinical utility of the BPSConnect protocol as a comprehensive bio-psycho-social screening tool for children with autism spectrum disorder (ASD), as well as to explore correlations across biological, psychological, and social dimensions.

**Methods:** A cross-sectional pilot study was conducted at e-TherapyKids, Barcelona (November 2024–June 2025). Twenty-five children with ASD, aged 4 to 8 years, diagnosed using the ADOS-2, were included. The protocol integrated checklists by dimension (biological, psychological, social), validated scales (SRS-2, Vineland-3, SSP2/SP2, PSI-SF, FES), and an exploratory hair epigenetic analysis (Cell Wellbeing). Data were pseudonymized and analyzed descriptively. Correlations between alarm indices and psychometric outcomes were explored using Spearman’s rho.

**Results:** Feasibility was high (completion rate >90%, mean administration time <60 minutes per dimension). Prevalence of alarm signals indicated frequent biological imbalances (64% vitamin D deficiency, 56% iron deficiency, 72% elevated exposure to electromagnetic fields), marked psychological difficulties (84% sensory processing, 68% social communication), and high social stressors (84% parental stress, 84% lack of school support). Exploratory correlations identified: vitamin D deficiency with socio-communicative impairment (ρ = 0.41), iron deficiency with language difficulties (ρ = 0.33) and attention/executive functions (ρ = 0.33), and parental stress with poorer adaptive functioning (ρ = –0.38) and behavioral dysfunction (ρ = 0.35).

**Conclusions:** The BPSConnect protocol proved feasible and clinically relevant in a real-world setting, allowing integrated detection of functional patterns across biological, psychological, and social domains. Findings suggest plausible associations between nutritional deficiencies, family stress, and neurodevelopmental outcomes. Although causality cannot be inferred due to the pilot design and the non-quantitative nature of biomarkers, results provide essential operational parameters for the multicenter NeuroEpigenCare study, which will incorporate standardized biomarkers, psychometric validation, and longitudinal follow-up.

## 1. Study: Bio-psycho-social correlation in children with ASD: preliminary results of the pilot application of the BPSConnect model

### 1.3 Ethics Committee Review

Ethics Committee for Research of e-TherapyKids (Ref. BPS-TEA/2024-2025-11). Constituted for the project, in accordance with the Helsinki Declaration and EU data protection regulations.

### 1.4 General Objective

To evaluate the feasibility and preliminary clinical utility of the BPSConnect instrument for comprehensive bio-psycho-social screening in ASD and to identify associations across dimensions.

### 1.5 Specific Objectives

- To determine the applicability of the protocol (completeness, time).
- To estimate the prevalence of alarm signals by dimension.
- To explore correlations between biological variables (including hair epigenetic test), psychological and social variables, and their relationship with performance and school environment.

### 1.6 Scientific and Clinical Rationale

#### Biological (relevant examples in ASD)

micronutrient deficiencies (vitamin D, iron), functional gastrointestinal dysfunction, allergies/intolerances, sleep disturbances, environmental exposures (e.g., EMFs).

#### Psychological/Behavioral and Language

socio-communicative difficulties, sensory processing disorders, repetitive behaviors/emotional regulation, expressive/receptive language alterations, and motor development.

#### Social

parental stress, caregiver burden, educational barriers (lack of methodological adaptation and specialized support), family climate, and support networks. The integration of these comorbidities allows for the detection of functional patterns and guides referrals and early interventions.

### 1.7 Theoretical Framework

#### 1.7.1 Foundation of the Bio-psycho-social Model

The understanding of autism spectrum disorders (ASD) has undergone a profound transformation in recent decades, shifting from reductionist models focused solely on behavioral or neuropsychological dimensions towards an integrative approach that considers multiple determinants of health and development. The bio-psycho-social (BPS) model, originally formulated by Engel in the 1970s and subsequently applied transversally in various fields of health, offers an interpretive framework particularly well-suited to address the complexity of ASD. This approach recognizes that the core symptoms of autism (socio-communicative alterations and repetitive behaviors) do not present in isolation but interact dynamically with medical conditions, psychological factors, and social contexts that together modulate clinical expression and functional prognosis (von Elm et al., 2007; World Medical Association, 2013).

In this sense, traditional diagnostic models based exclusively on behavioral criteria, while useful for nosological classification, are insufficient to explain the clinical heterogeneity and diverse developmental trajectories observed in children with ASD. Scientific literature increasingly supports the need for transversal frameworks that integrate biomedical, psychological, and social data to guide early, individualized, evidence-based interventions (Frye et al., 2020; Qi et al., 2023).

#### 1.7.2 Biological Evidence and Comorbidities

The biological dimension of ASD is characterized by a high frequency of medical comorbidities that significantly affect quality of life and functionality. Among these, nutritional deficiencies, metabolic alterations, and environmental epigenetic factors stand out. Numerous studies have documented deficiencies in vitamin D and iron as recurrent findings in children with ASD, with these parameters associated with greater severity of socio-communicative symptoms and increased risk of cognitive and behavioral difficulties (Bener et al., 2014; Bener et al., 2017; Cannell & Grant, 2013).

A particularly relevant line of research addresses food selectivity, a frequent phenomenon in children with ASD, characterized by marked dietary restriction, aversion to certain textures, colors, or food temperatures, and preference for a limited range of foods. Recent studies show that food selectivity is not a mere behavioral trait but a direct risk factor for the development of clinically relevant nutritional deficiencies. Sharp et al. (2013) reported that children with ASD are three to five times more likely to suffer from micronutrient deficiencies such as vitamin D, B12, folate, iron, and zinc compared with their typically developing peers, and that these deficits correlate with the exacerbation of behavioral and sensory problems. Additional evidence by Bandini et al. (2017) confirms that food neophobia and selectivity are associated with reduced intake of fruits, vegetables, and proteins, thereby aggravating the metabolic vulnerability of this population.

The relationship between nutritional deficiencies and ASD symptoms has been widely discussed in the literature. For example, low vitamin D levels have been associated with greater severity on the SRS-2 scale, indicating a link between nutritional status and socio-communicative functioning (Fernell et al., 2015). Similarly, low iron and ferritin levels have been associated with sleep disturbances, attention deficits, and executive dysfunction (Bener et al., 2017; Schmidt & Tancredi, 2021). In this line, Frye et al. (2020) argue that micronutrient deficiencies and alterations in metabolic biomarkers could represent a “second biological hit” that intensifies the severity of core autism symptoms.

On the other hand, epigenetic and environmental factors have gained importance as modulators of ASD phenotypic expression. Recent research suggests that environmental exposures such as chemical pollutants, heavy metals, or non-ionizing electromagnetic radiation can induce epigenetic changes affecting gene regulation involved in neurodevelopment (Gialloreti et al., 2022; Qi et al., 2023). Although the evidence is still emerging, these findings reinforce the need to integrate exploratory epigenetic indicators into assessment protocols—not as definitive diagnostics, but as potential risk signals that justify preventive interventions.

#### 1.7.3 Psychological and Behavioral Dimension

In the psychological dimension, ASD is primarily defined by alterations in socio-communication and the presence of restricted and repetitive behavior patterns (Lord et al., 2012). However, contemporary approaches emphasize that these characteristics must be analyzed in relation to other areas of psychological functioning, such as sensory processing, emotional regulation, and language development.

Sensory processing has emerged as an area of particular interest in recent literature. Dunn et al. (2016) and Tomchek et al. (2018) have shown that up to 80% of children with ASD present patterns of hyper- or hyposensitivity that directly affect their participation in school and family environments. These alterations have been correlated with greater difficulties in emotional regulation and with an increase in disruptive behaviors, reinforcing the importance of evaluating this dimension as an integral part of functional diagnosis.

Language constitutes another central axis. Longitudinal studies have demonstrated that alterations in expressive and receptive language predict the developmental trajectory of social and cognitive competencies in children with ASD (Tager-Flusberg, 2016). Moreover, recent research has linked the severity of language delay with nutritional deficiencies, particularly of iron and vitamin D, further reinforcing the interconnection between biological and psychological dimensions (Bener et al., 2017; Frye et al., 2020).

Emotional and behavioral regulation, frequently compromised in children with ASD, is consistently associated with both biological factors (micronutrient deficiencies, metabolic dysfunctions) and social ones (parental stress, lack of school support). The study by Mazefsky et al. (2013) shows that difficulties in emotional regulation not only intensify challenging behaviors but also mediate the relationship between sensory deficits and behavioral problems, constituting a key point for interdisciplinary interventions.

#### 1.7.4 Social and Family Dimension

The social and family dimension is decisive in shaping the clinical picture of ASD and in determining the response to interventions. Literature has robustly documented that parents of children with ASD exhibit higher levels of stress, anxiety, and depression than parents of typically developing children or those with other neurodevelopmental conditions (Hayes & Watson, 2013; Rivard et al., 2014). Parental stress is not only a consequence of the diagnosis but is directly associated with symptom severity in the child and the emergence of additional behavioral difficulties (Karhu et al., 2022; Martínez-González et al., 2020).

Several studies have identified that elevated parental stress correlates with a higher prevalence of emotional and behavioral regulation problems in children, generating a negative feedback loop: higher caregiver stress reduces containment capacity, which in turn exacerbates the child’s symptoms (Rivard et al., 2014; Karhu et al., 2022). Likewise, the absence of educational support, such as lack of curricular adaptations, insufficient specialized staff, or weak school-family coordination, exacerbates parental burden and limits the child’s learning opportunities (Moos, 1994; Sparrow et al., 2016).

In the school setting, literature has documented that the lack of methodological adaptation and specialized support constitutes a critical factor that amplifies children’s psychological difficulties and increases family overload (Karhu et al., 2022). Inclusive education policies, when effectively applied, not only improve access to resources and school participation but also act as protective factors against the chronicity of parental stress and the emergence of secondary behavioral problems (Rivard et al., 2014; Martínez-González et al., 2020). In this sense, the bio-psycho-social framework cannot be understood without recognizing the relevance of the family-school interaction, which constitutes a strategic axis for early intervention and improved functional prognosis. Indeed, the European Agenda for Educational Inclusion and UNESCO’s normative framework on inclusive education have pointed out that the success of ASD care depends as much on the accessibility of environments as on the training of education professionals.

This evidence underscores that the social dimension cannot be understood as a secondary factor but as an active modulator of the clinical course of ASD, whose influence must be systematically assessed in any diagnostic or intervention protocol.

#### 1.7.5 Interaction Across Dimensions and the BPSConnect Framework

Integrating the three dimensions described biological, psychological, and social is essential for advancing towards a functional rather than merely classificatory diagnosis. Recent findings show that biological deficits such as vitamin D or iron deficiency not only affect physiological processes but also have a direct impact on cognition, communication, and behavior (Bener et al., 2014; Frye et al., 2020). Similarly, social conditions, parental stress, educational barriers, modulate the clinical expression of psychological symptoms, reinforcing or attenuating challenging behaviors and socio-communicative difficulties (Hayes & Watson, 2013; Karhu et al., 2022).

The BPSConnect model situates itself precisely at this intersection of interactions, proposing a systematic protocol that identifies correlations across dimensions and prioritizes intervention objectives based on functional patterns. The preliminary results of the pilot reinforce this relevance: the observed correlation between vitamin D deficiency and socio-communicative dysfunction, between parental stress and reduced adaptive participation, or between low iron and difficulties in language and attention are not isolated findings, but empirical confirmations of what has already been highlighted in international literature. Thus, the model offers a practical operationalization of the bio-psycho-social approach, transforming a theoretical framework into a clinically applicable tool in real-world settings.

## 2. Conceptual Framework and Hypotheses

### 2.1 Three-Dimensional Connected Validation Model

The BPSConnect model is conceived as an innovative proposal that addresses the need to overcome fragmented approaches in the assessment of autism spectrum disorders, integrating in a structured way three fundamental dimensions of human development: the biological, the psychological, and the social. The logic underpinning this model rests on the premise that the observable symptoms in a child with ASD are not exclusively the result of neurological alterations, but rather emerge from the complex interaction between biological predispositions, behavioral manifestations, and contextual conditions. From this perspective, the protocol combines structured screening instruments, validated scales, and an exploratory analysis of epigenetic biomarkers in order to construct a comprehensive framework for assessment.

The operational core of the model is organized around checklists designed for each dimension, which allow the systematic and standardized identification of alarm signals. These checklists are complemented by internationally recognized scales, such as the SRS-2 for socio-communication, the Vineland-3 for adaptive behavior, the SSP2/SP2 for sensory profiles, the PSI-SF for parental stress, and the FES for family climate. The choice of these instruments is not arbitrary but responds to criteria of psychometric validity, availability in Spanish, and their pertinence in capturing the key phenomena that modulate the child’s functioning. The incorporation of the exploratory hair epigenetic analysis from Cell Wellbeing, although non-diagnostic, adds a novel component that enables the exploration of correlations between nutritional deficiencies or environmental exposures and psychological and social manifestations.

The ultimate aim of this connected three-dimensional model is not merely to accumulate data, but to generate functional correlations that can be used to prioritize intervention objectives. Unlike traditional protocols, which tend to provide isolated categorical diagnoses, BPSConnect seeks to establish relationships across dimensions that explain why a child manifests certain difficulties in a given context, and how these difficulties may be modified through interventions directed not only at the child but also at the family and school environment.

### 2.2 Hypotheses

The formulation of hypotheses in this pilot responds to the need to explore plausible associations across domains of different nature, consistent with the scientific evidence accumulated in recent years. The first hypothesis posits that a higher burden of biological signals, particularly nutritional deficiencies such as low levels of vitamin D and iron, will be associated with poorer performance in socio-communicative, language, and executive function domains, as well as with a greater prevalence of sensory alterations, both hyper- and hyporeactivity. This hypothesis is based on prior research describing the relevance of micronutrients in the modulation of neurotransmitters and neuronal plasticity, suggesting a direct impact on communication and sensory regulation.

The second hypothesis proposes that elevated levels of parental stress, assessed through the PSI-SF, will be associated with lower adaptive participation in the Vineland-3, with an increase in behavioral difficulties, and with poorer school performance, reflected in the need for specialized support and in a lower adequacy of the classroom to the child’s needs. This hypothesis is grounded in studies documenting the bidirectional relationship between caregiver stress and the severity of the child’s symptoms, forming a vicious cycle in which behavioral difficulties exacerbate family distress, which in turn limits the child’s capacity to function in adaptive contexts.

The third hypothesis postulates that insufficient school supports and inadequate methodological adaptations are related to greater severity in socio-communicative and behavioral domains. This proposition responds to evidence indicating that the school is not a mere passive setting for symptom observation, but rather a space that can amplify or attenuate the expression of difficulties depending on the quality of supports and the pedagogical flexibility implemented.

### 2.3 Endpoints

In coherence with the exploratory nature of the pilot, primary and secondary endpoints were defined. The primary endpoint focused on the feasibility of the protocol, establishing as success criteria a completion rate above 90%, a total administration time of less than 60 minutes per dimension, and the absence of adverse events derived from the evaluation process. The definition of these parameters responds to the need to demonstrate that the protocol is not only scientifically sound but also operationally sustainable.

Secondary endpoints included the estimation of prevalence of alarm signals by dimension, the calculation of Spearman correlations between alarm indices and validated scales, and the exploration of specific associations between nutritional deficiencies indicated by hair epigenetic biomarkers and key variables such as communication, language, attention, executive functions, and sensory processing. These endpoints allow progress in validating the relevance of the model and in identifying relationships that justify larger confirmatory studies.

## 3. Methodology (STROBE)

### 3.1 Design

The study was designed as a cross-sectional observational pilot, conducted in a real-world clinical setting. The choice of this design reflects the intention to explore correlations and patterns of association without attempting to establish causal relationships, in line with methodological guidelines for exploratory studies that precede confirmatory clinical trials.

### 3.2 Population, Recruitment, and Sample

The target population consisted of children with a confirmed diagnosis of ASD according to DSM-5 criteria and verification via ADOS-2, aged between 4 and 8 years. Recruitment took place at the e-TherapyKids clinic in Barcelona, during the period from November 2024 to June 2025. A convenience sample of 25 participants was selected, considering inclusion criteria related to age, diagnosis, and availability of parental consent, as well as exclusion criteria associated with severe neurometabolic disorders, refractory epilepsy, or refusal to provide consent. The selection flow was documented in an eligibility diagram that reflected the stages of invitation, inclusion, and analysis, in accordance with STROBE recommendations.

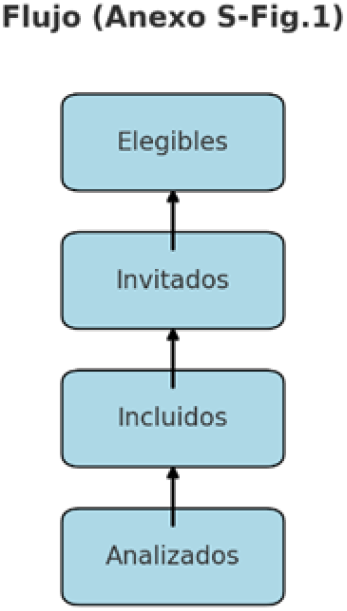

### 3.3 Procedure

Each participant was evaluated across three dimensions biological, psychological, and social by an interdisciplinary team composed of specialists in psychology, speech therapy, physiotherapy, and clinical nutrition. The assessments were recorded in an electronic system designed to ensure data pseudonymization, in compliance with current data protection regulations in the European Union. This procedure ensured homogeneity in data collection and minimized the risk of biases arising from variability in records.

### 3.4 Dimension-Specific Checklists and Thresholds

Dimension-specific checklists were applied to compute the percentage of alarm items in defined subdomains, establishing categorization thresholds ranging from minimal (<25%), mild (≥25%), moderate (≥50%), to severe (≥75%). In the biological dimension, the subdomains evaluated included gastrointestinal, neurological, ENT, ophthalmological, allergies and intolerances, as well as hair epigenetic biomarkers and exposure to electromagnetic fields. In the psychological dimension, the subdomains encompassed language and communication, social skills, emotional-behavioral regulation, sensory processing, attention and executive functions, and motor development. Finally, in the social dimension, parental stress, family climate, support network, classroom adequacy, curricular adaptation, and specialized support were analyzed. This scheme allowed for capturing the complexity of the child’s functioning across multiple domains and generating differentiated risk profiles.

### 3.5 Validated Scales

The validity of the protocol was reinforced by the application of standardized scales recognized in international clinical practice. The ADOS-2 was used to confirm ASD diagnosis. The SRS-2 assessed the severity of socio-communicative impairment. The Vineland-3 was employed to evaluate adaptive behavior in its domains of communication, socialization, and daily living skills. The SSP2/SP2 provided a detailed profile of sensory processing. The PSI-SF offered information on levels of parental stress, while the FES allowed for analysis of family climate and cohesion/conflict patterns. All scales were administered in their Spanish versions, ensuring cultural validity and comprehension of the measures.

### 3.6 Hair Epigenetic Biomarkers

The biological analysis relied on the use of non-invasive hair technology from Cell Wellbeing, through the S-Drive device. This procedure, employed for preventive and non-diagnostic purposes, enabled the identification of alarm signals related to nutritional deficiencies such as vitamin D and iron, as well as environmental exposures including electromagnetic fields and heavy metals. The methodological decision to incorporate this technique stemmed from the interest in exploring low-risk, low-cost, and highly acceptable tools in pediatric ASD populations, where invasive extractions are often complex. Minimization of clinical risk and the high tolerance of children represented additional justifications for its inclusion in the pilot. Nevertheless, it is acknowledged that the absence of standardized units limits diagnostic inference. In the future validation phase, it is planned to incorporate quantitative reference methodologies such as ICP-MS for trace elements, LC-MS/MS for serum vitamin D, and dosimetry instruments for the evaluation of electromagnetic exposures, thereby allowing for more robust triangulation of data.

### 3.7 Data, Security, and Confidentiality

Data management was carried out under strict protocols of security and confidentiality, in compliance with the General Data Protection Regulation (GDPR) and the Spanish Organic Law on Data Protection and Digital Rights (LOPDGDD). Records were pseudonymized, stored on encrypted servers located within the European Union, and accessible only to authorized profiles. The data retention period was set at five years, in accordance with applicable regulations for observational clinical studies. Additionally, data quality control measures were implemented, including inter-rater review to ensure coherence across professionals, double data entry to reduce transcription errors, and complete-case analysis in handling missing values, avoiding imputations that could introduce bias.

### 3.8 Statistical Plan

The statistical analysis was structured into two phases. In the first, descriptive in nature, means and standard deviations were calculated for normally distributed continuous variables, medians and interquartile ranges for non-parametric variables, and proportions with 95% confidence intervals for categorical variables, using Wilson’s method. In the second, exploratory phase, Spearman correlations were calculated between alarm indices obtained from the checklists and scores from validated scales, as well as associations with hair epigenetic markers. Additionally, simple linear models adjusted for age and sex were applied in order to describe trends without aiming for confirmatory inference. No imputation of missing data was performed, and the analysis was conducted using complete cases, in line with the exploratory nature of the pilot study.

## 4. Ethical Aspects

This study adheres to the international ethical principles governing research with human subjects, particularly the Declaration of Helsinki (World Medical Association, 2013), and complies with European and Spanish regulations on personal data protection (GDPR and LOPDGDD). From its design, the research was considered to involve minimal risk for participants, as clinical and social assessments were conducted through interviews, standardized scales, and non-invasive procedures. At no point were drugs administered or interventions carried out that could cause direct physical or psychological harm to the children. The hair epigenetic analysis, employed for exploratory purposes, was based on hair sample collection—a procedure with no medical risks and negligible discomfort for participants. This assessment of minimal risk was confirmed by the e-TherapyKids Research Ethics Committee (REC), which reviewed the documentation and authorized the study under reference BPS-TEA/2024-2025-11.

The potential benefits of participation were considered significant in relation to the minimal burden of risks. Among these were the opportunity to obtain an integrated detection of alarm signals across the biological, psychological, and social dimensions, with early clinical and educational guidance that could be useful for families in decision-making regarding treatments and school supports. While the study did not aim to provide a clinical diagnosis or prescribe direct interventions, the generated reports could serve as preliminary guidance for caregivers to discuss with their physicians, therapists, or educators, thereby promoting a more coordinated approach to the child’s needs. From a bioethical perspective, this balance between low risk and potential benefit fully justifies the implementation of the pilot study.

Regarding informed consent, a standardized process was designed to ensure that families understood the nature of the study, its objectives, the voluntary nature of participation, and the possibility of withdrawal at any time without any negative consequences for the care received at the clinic. For this purpose, specific documents (Annexes S-CI1 and S-CI2) were prepared in clear, accessible, jargon-free language, which were reviewed with parents or legal guardians prior to signing. During this process, dedicated time was provided to address questions, and it was ensured that the decision to participate was made freely and in an informed manner. In line with international ethical recommendations, any form of coercion was avoided, presenting the study as a voluntary option that did not condition access to the clinic’s regular services.

Particular attention was given to the vulnerable condition of children with ASD and their families. To ensure a respectful and protective approach, interviewers received specific training in communication techniques adapted to neurodiversity and in strategies to support caregivers who might experience emotional overload during the interview process. Support mechanisms were also in place so that if a parent expressed distress during evaluations, emotional support was offered along with the option to interrupt or reschedule the session. This sensitivity to vulnerability is not only part of the ethical obligations of the researcher but also a fundamental pillar of e-TherapyKids’ intervention philosophy, centered on the comprehensive care of the child and family.

Confidentiality and data security constituted another priority within the ethical dimension of the study. Pseudonymization mechanisms were implemented from the moment of data collection, ensuring that participant identity could not be linked to the collected data. Data storage was conducted on encrypted servers located in the European Union, with access restricted only to authorized members of the research team, in accordance with the principle of access minimization. Data will be retained for five years, in compliance with applicable regulations, and subsequently deleted securely, avoiding any risk of improper disclosure.

### 4.1 Justification of the Pilot

The implementation of a pilot study such as the one presented here responds to a highly relevant ethical, scientific, and methodological justification. From an ethical perspective, it would not be acceptable to immediately implement a complex protocol such as BPSConnect in a large-scale study without first demonstrating its safety, applicability, and acceptability by families. The pilot design allows for refinement of instruments, detection of potential difficulties in administration, and adjustment of procedures before exposing a larger number of participants. In this way, risks are minimized and future trials, such as the projected NeuroEpigenCare, can be conducted under safer and more efficient conditions.

From a methodological standpoint, the pilot is essential for estimating the variance of key outcome measures and, consequently, calculating the sample size required for confirmatory studies with sufficient statistical power. The experience gathered in this preliminary phase is also crucial for anticipating the logistical resources that must be mobilized in a multicenter trial, as well as for establishing realistic parameters regarding administration times, family burden, and instrument completion rates. In short, the justification for the pilot lies not only in the scientific interest of exploring preliminary associations but also in the ethical obligation to proceed gradually, ensuring that each step is based on evidence of feasibility and respect for participants.

Furthermore, this study acquires an innovative and pioneering character in the European context, as it represents the first pilot to simultaneously integrate exploratory epigenetic biomarkers, standardized psychometric scales, and social factors into a single screening and intervention protocol. This methodological convergence broadens the scope of clinical and scientific interpretation, by allowing integrated observation of how biological, psychological, and social factors interact in the symptomatic expression of children with ASD. At the same time, the approach responds to the growing international demand for translational research models that combine biomedical precision with sensitivity to family and school contexts.

The relevance of this approach is reinforced when preliminary findings are compared with previous multicenter experiences, which have demonstrated the influence of nutritional deficiencies, environmental factors, and parental overload on the developmental trajectories of children with ASD across different countries (Bener et al., 2017; Hayes & Watson, 2013; Gialloreti et al., 2022). Although the sample sizes of this pilot do not allow definitive conclusions, the consistency of the observed signals with international literature supports the model’s direction and suggests that BPSConnect may become a useful tool for generating comparable evidence in future European and intercontinental multicenter studies.

In this sense, the justification for the pilot not only responds to the need to establish methodological conditions for the NeuroEpigenCare trial but also to the commitment to position BPSConnect within an international network of neurodevelopment research. Thus, the present work lies at the intersection of clinical innovation, research ethics, and international projection, consolidating a pathway toward the validation of an integrated protocol that may transform clinical and educational practice in the management of ASD.

## 5. Results

### 5.1 Baseline Characteristics

**Table 1.**

5.1 Baseline Characteristics. Table 1
| Variable | Value |
| --- | --- |
| Age (years), mean ± SD | 6.2 ± 1.3 |
| Male sex, n (%) | 18 (72%) |
| Female sex, n (%) | 7 (28%) |
| Medical comorbidity, n (%) | 10 (40%) |
| Diagnosis confirmed with ADOS-2 | 25 (100%) |

### 5.2 Alarm Signals

#### Biological (hair epigenetic test)

**Table 2.**

5.2 Alarm Signals Biological (hair epigenetic test). Table 2
| Subdomain | Indicator | n/N (%) |
| --- | --- | --- |
| Deficiencies (hair, pilot) | Low Vitamin D | 16/25 (64%) |
|  | Low Iron | 14/25 (56%) |
|  | Low Magnesium | 7/25 (28%) |
|  | Low B1/B9 | 8/25 (32%) |
|  | Low Linoleic Acid | 3/25 (12%) |
| Exposure | High EMF | 18/25 (72%) |
|  | Heavy Metals | 9/25 (36%) |
| Systems (checklist) | Neurological | 9/25 (36%) |
|  | Gastrointestinal | 7/25 (28%) |
|  | ENT | 5/25 (20%) |
|  | Ophthalmological | 2/25 (8%) |
|  | Allergies/Intolerances | 3/25 (12%) |

**Fig. SR2.1.**
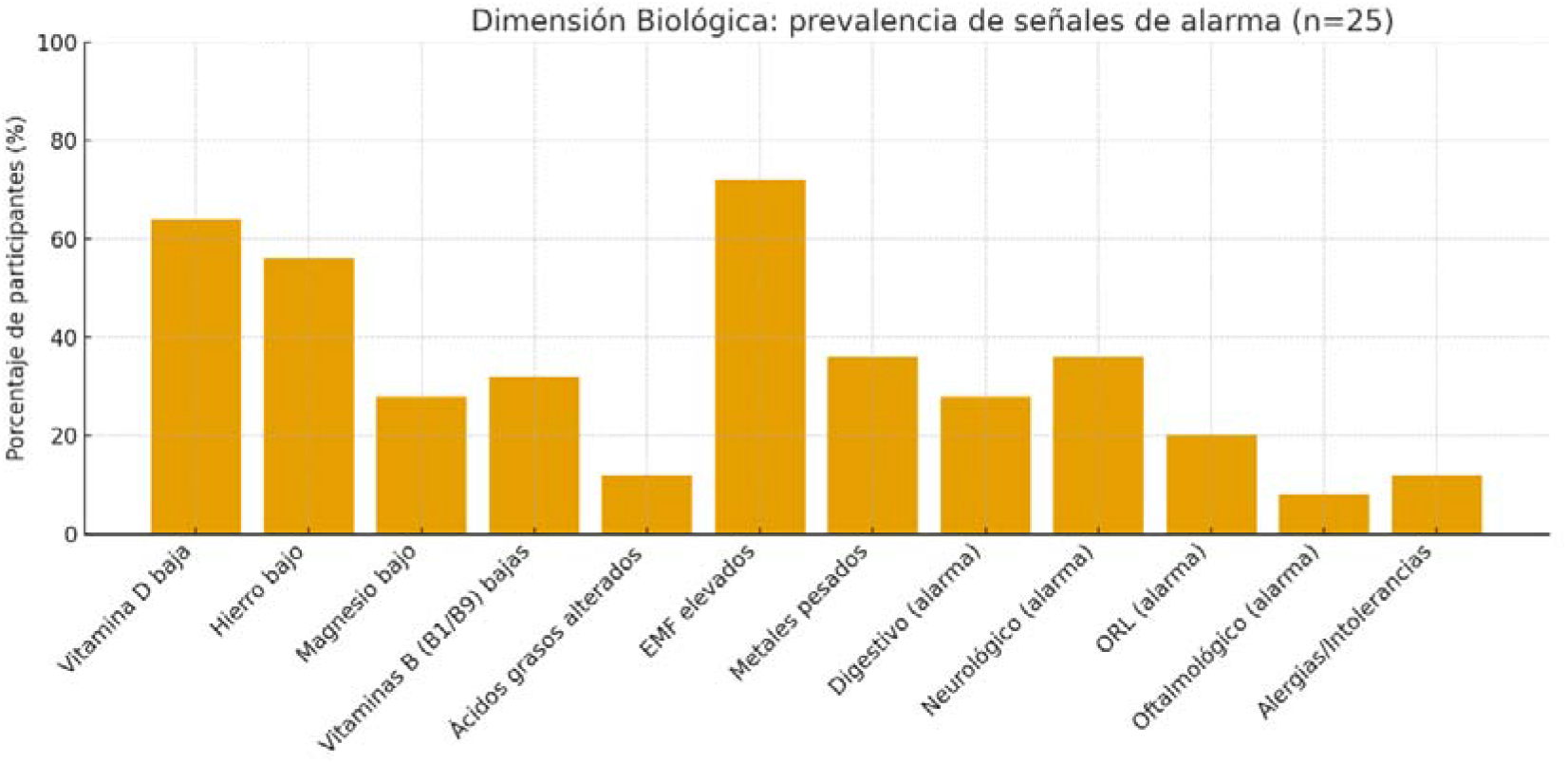
(Biological): Prevalence of biological alarm signals in the sample (n=25). Epigenetic “alarms” (VitD, Fe, Mg, B vitamins, fatty acids, EMF, metals) are interpreted as exploratory signals (preventive hair test). Clinical subdomains (gastrointestinal, neurological, ENT, ophthalmological, allergies/intolerances) derive from the BPSConnect checklist. Validity note: Hair-based measurements were non-quantitative (Cell Wellbeing platform, preventive focus). They are reported as exploratory signals and not as biochemical diagnoses.

### 5.3 Psychological Dimension (detail and threshold gradation)

**Table 3.**
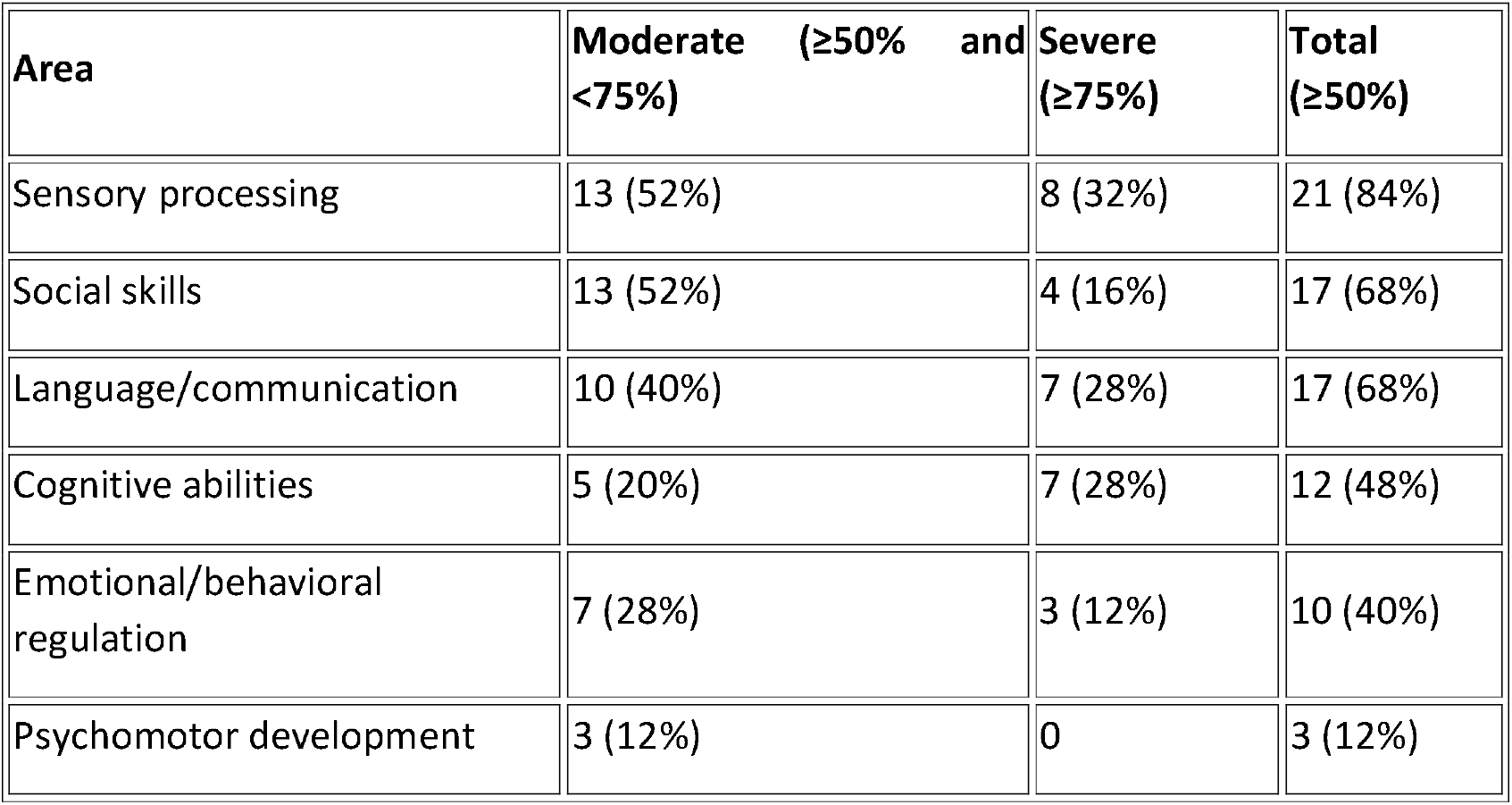

**Fig. SR2.2.**
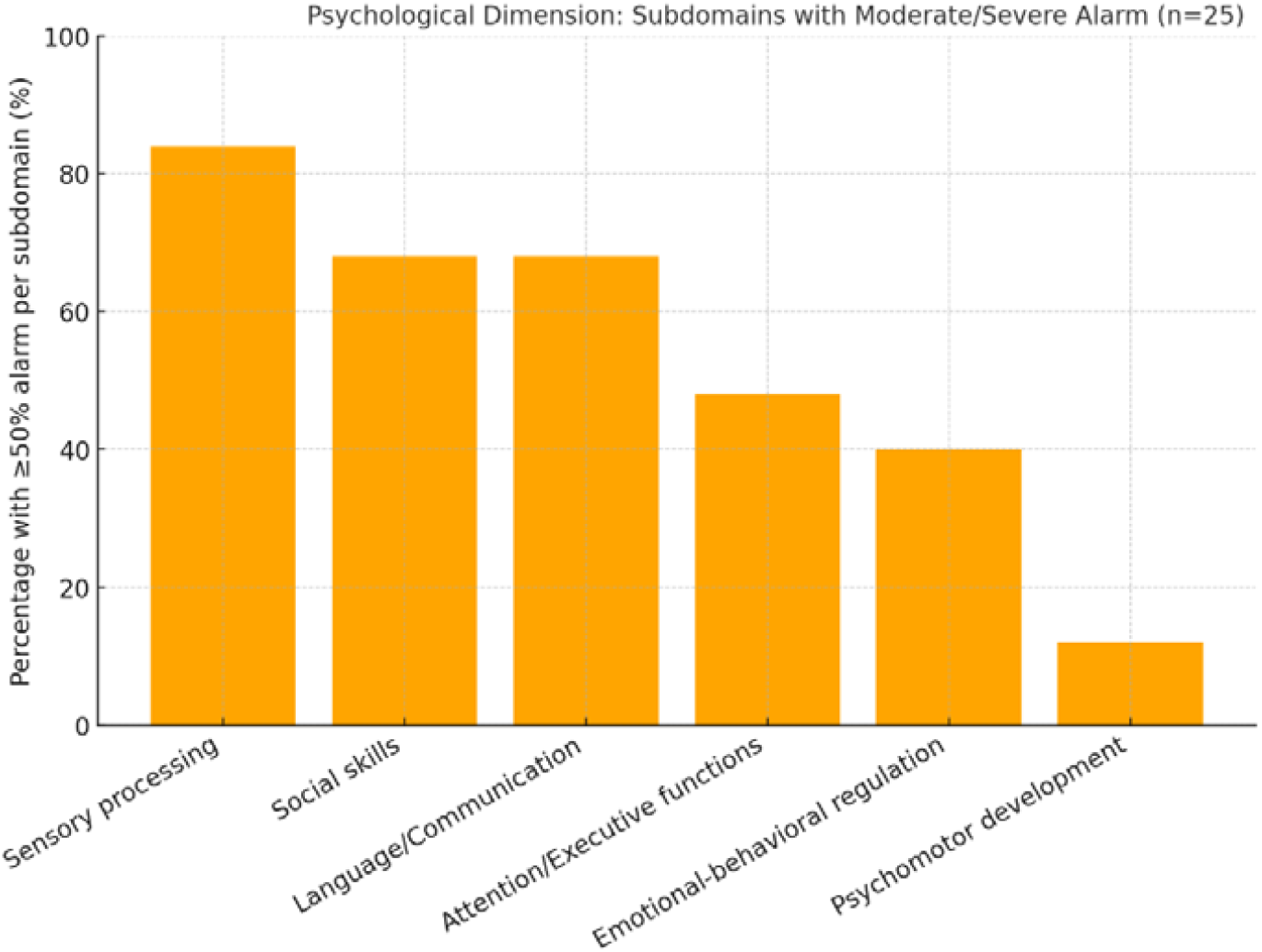
(Psychological): Percentage of participants with ≥50% of alarm items per subdomain (moderate/severe). Higher load observed in sensory processing and socio-communication. Results derive from age-specific checklists and are interpreted alongside validated scales (SRS-2, Vineland-3, SSP2/SP2).

### 5.4 Social Dimension (family and school)

**Table 4.**

5.4 Social Dimension (family and school). Table 4
| Subdomain | Indicator | n/N (%) |
| --- | --- | --- |
| Family | High parental stress | 21/25 (84%) |
| | Anxiety in $\geq 1$ caregiver | 20/25 (80%) |
| | Depression in $\geq 1$ caregiver | 6/25 (24%) |
|  | Poor intrafamilial communication | 15/25 (60%) |
|  | Dysfunctional separation | 5/25 (20%) |
|  | No support network | 1/25 (4%) |
| School | Lack of classroom adaptation | 11/25 (44%) |
|  | Lack of curricular/methodological adaptation | 10/25 (40%) |
|  | Lack of specialized classroom support | 21/25 (84%) |

**Fig. SR2.3.**
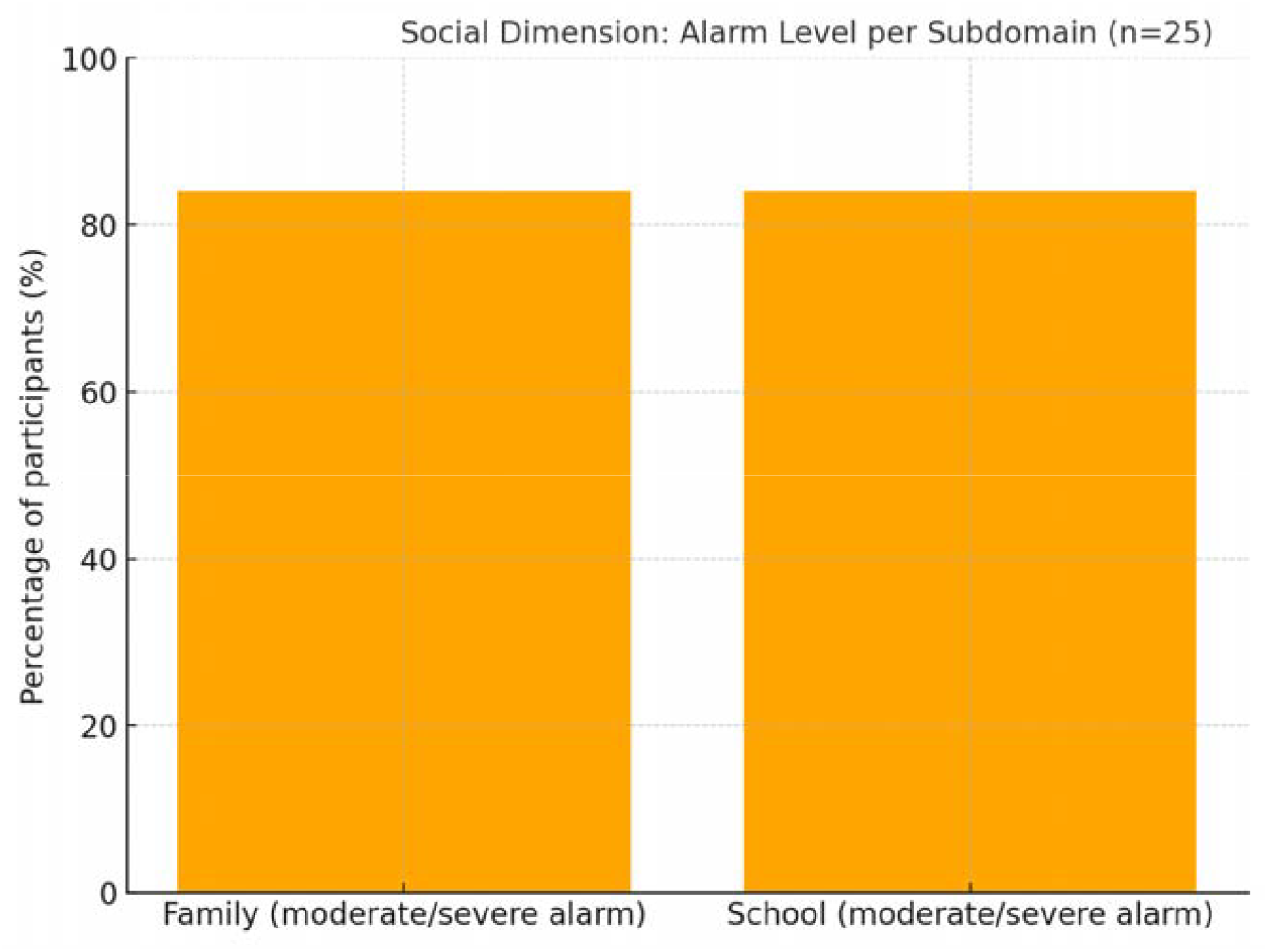
(Social): Proportion of cases with moderate/severe alarm in the family context (e.g., high parental stress) and school context (e.g., insufficient supports). Both subdomains show high alarm prevalence in the pilot.

### 5.5 Exploratory Correlations (Spearman) and Clinical Signal

**Table 5.**
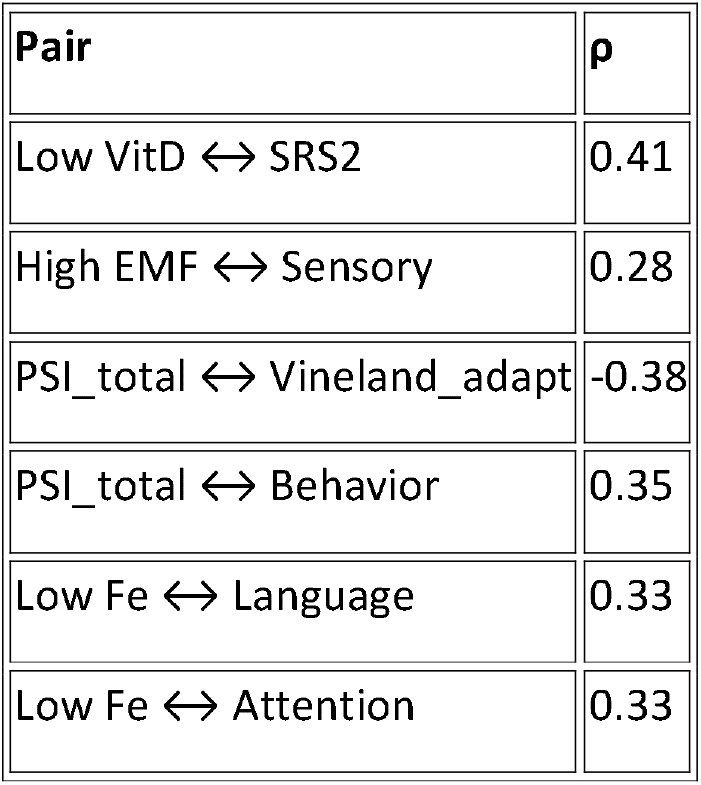

**Fig. SR2.4.**
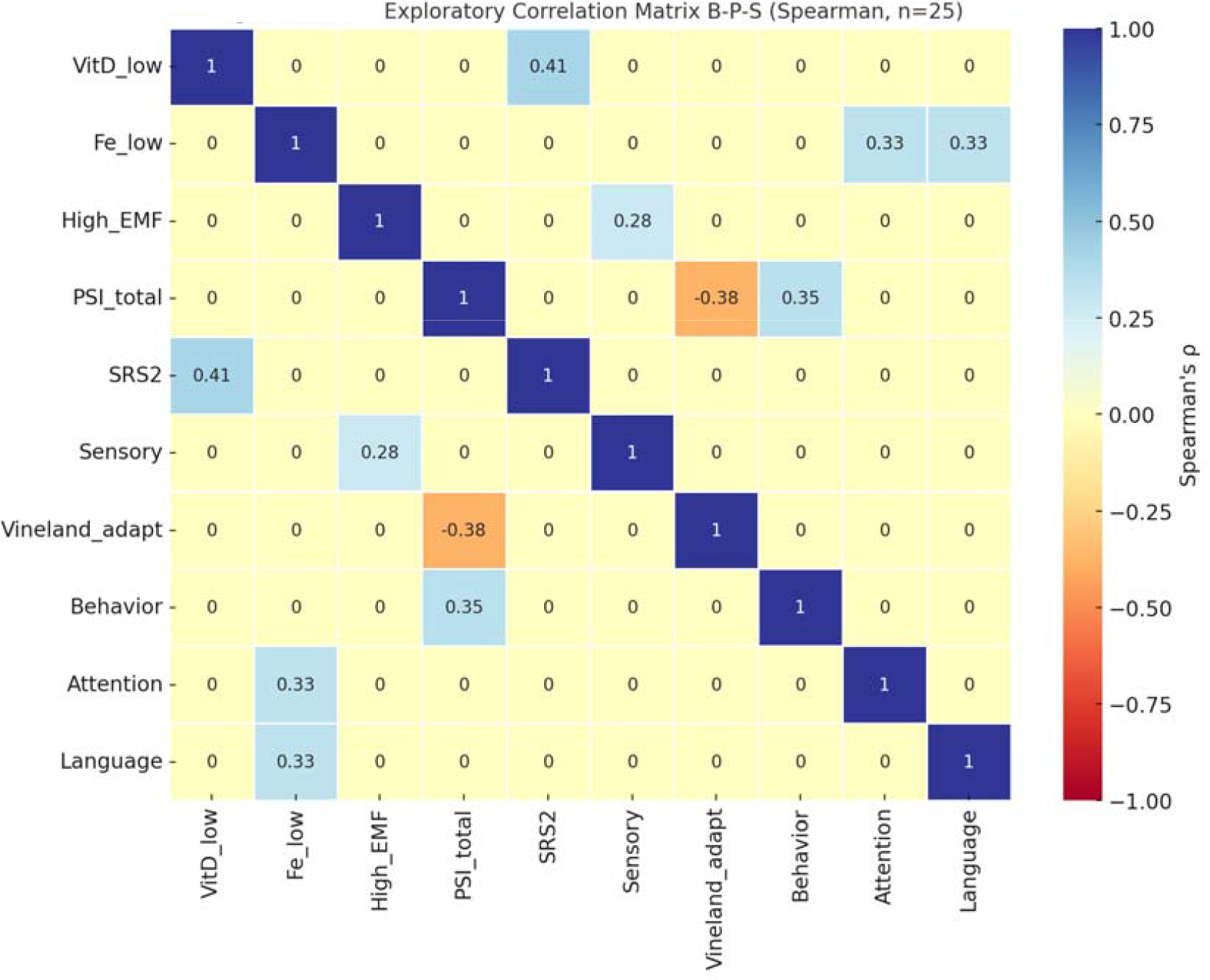
(Spearman correlation pairs): Exploratory correlation matrix of B-P-S (Spearman, n=25). The matrix represents exploratory correlations between biological variables (Low VitD, Low Fe, High EMF), social variables (PSI_total), and psychological/functional variables (SRS2, Sensory, Vineland_adapt, Behavior, Attention, Language). Most notable associations included: low vitamin D with socio-communicative impairment (ρ=0.41), low iron with language difficulties (ρ=0.33) and attention/executive deficits (ρ=0.33), and parental stress with reduced adaptive functioning (ρ=−0.38) and behavioral dysfunction (ρ=0.35). A positive correlation was also observed between EMF exposure and sensory processing alterations (ρ=0.28).

**Fig. SR2.5.**
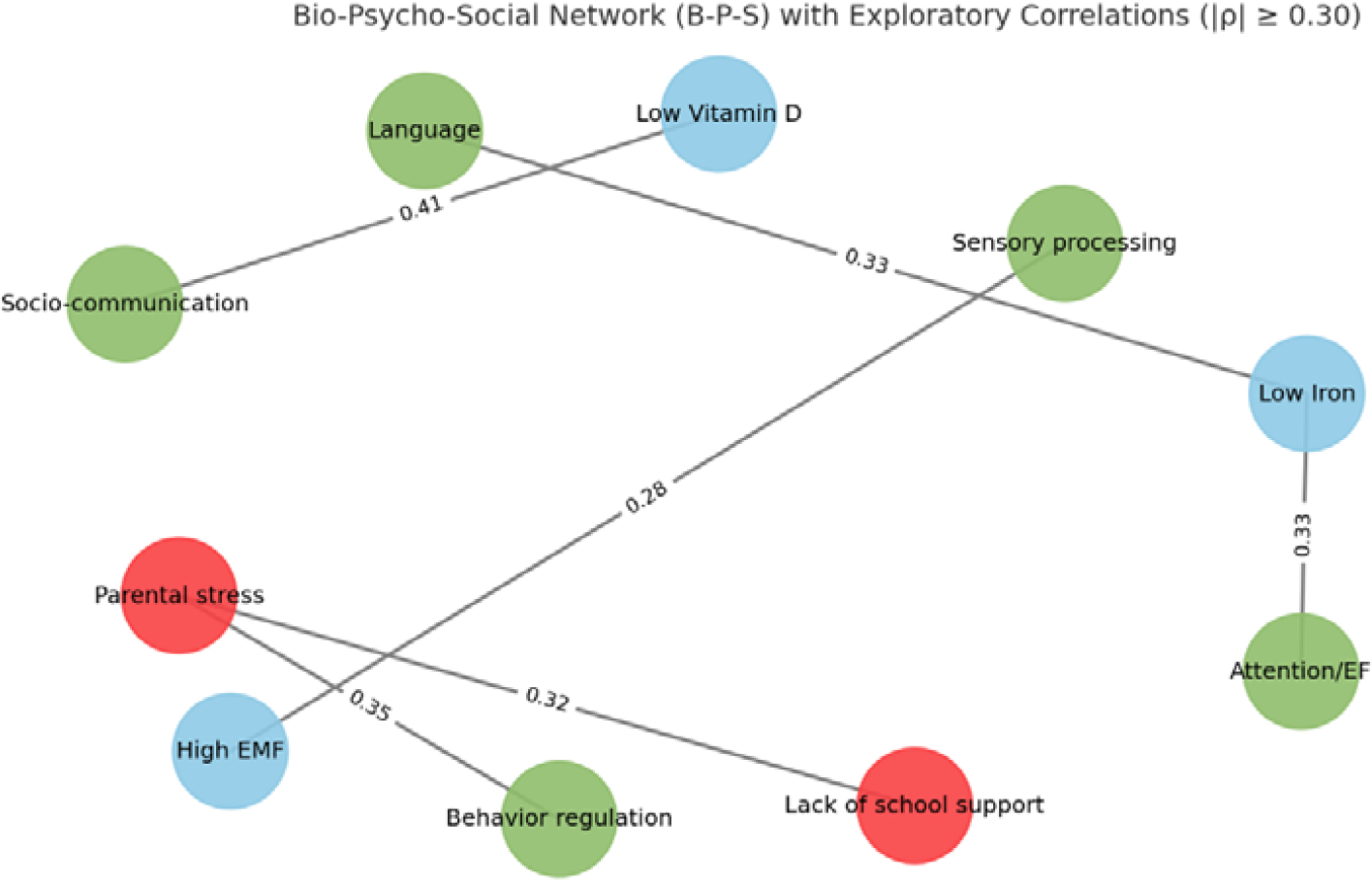
Bio-psycho-social (B-P-S) network with exploratory correlations (|ρ| ≥ 0.30): The diagram shows the most relevant associations among biological variables (blue nodes), social variables (red nodes), and psychological/functional variables (green nodes) in the pilot sample. Connections represent Spearman correlations equal to or greater than 0.30 in absolute value, with magnitude indicated on each edge. Highlights include the relationship between vitamin D deficiency and socio-communication difficulties (ρ=0.41), low iron with language (ρ=0.33) and attention/executive functions (ρ=0.33), as well as the association between parental stress and behavioral regulation problems (ρ=0.35) and lack of school supports (ρ=0.32). A trend was also noted between EMF exposure and sensory processing alterations (ρ=0.28).

The exploratory correlation analysis identified associations of interest among biological, psychological, and social variables that, while requiring cautious interpretation due to the small sample size, provide clinically and scientifically plausible signals that justify further validation studies. The most notable association was observed between vitamin D deficiency and socio-communicative dysfunction measured by the SRS-2, with a moderate positive correlation (ρ≈0.41). This finding suggests that children presenting indicators of low vitamin D availability also tended to manifest greater difficulties in social interaction and communication, consistent with prior research linking vitamin D to neuronal plasticity, neurotransmitter synthesis, and immune regulation, all relevant to neurodevelopment (Bener et al., 2014; Cannell & Grant, 2013).

In the social and family domain, a negative correlation was detected between parental stress level, measured with the PSI-SF, and adaptive participation recorded in the Vineland-3 (ρ≈−0.38). This association indicates that higher caregiver stress was related to lower levels of adaptive functioning in the child’s daily life. This finding is clinically relevant, as it underscores the influence of the family context on the expression of adaptive skills and highlights the need to consider caregiver well-being as a moderating variable in the developmental trajectory of children with ASD (Hayes & Watson, 2013; Martínez-González et al., 2020).

Furthermore, parental stress showed positive correlations in the range of 0.30 to 0.35 with the presence of behavioral regulation difficulties. This result aligns with literature documenting the bidirectional relationship between caregiver distress and intensification of behavioral problems in children, generating a negative feedback loop in which both phenomena reinforce each other (Rivard et al., 2014). In parallel, a positive correlation of similar magnitude was observed between parental stress and school outcomes, particularly the need for additional support and poorer classroom adaptation. These findings reinforce the notion that family stress not only impacts the home environment but also projects onto the child’s educational experience, contributing to more limited academic performance and greater reliance on specialized resources.

In the biological domain, iron deficiency, present in over 50% of the sample, was positively associated with greater language and communication difficulties, both in the SRS-2 and in the Vineland-3 communication subscale, with correlation coefficients in the 0.30–0.35 range. This clinical signal suggests that iron deficiency, an essential micronutrient for myelination and dopaminergic synthesis, may be linked to alterations in neurocognitive domains central to communication. Similarly, a positive correlation of comparable magnitude was identified between iron deficiency and difficulties in attention and executive functions, consistent with evidence linking iron to cerebral energy metabolism and performance in tasks requiring inhibitory control and cognitive flexibility (Frye et al., 2020).

Finally, in the environmental domain, a positive trend (ρ≈0.28) was observed between high exposure to electromagnetic fields (EMF) and sensory processing alterations assessed with the SSP2/SP2 profiles. Although this correlation does not reach a strong magnitude threshold, it constitutes a preliminary signal warranting further exploration of the potential interaction between environmental exposures and sensory sensitivity in children with ASD, consistent with recent reviews emphasizing the cumulative impact of environmental factors on neurodevelopment (Gialloreti et al., 2022).

Taken together, these correlations, although exploratory, exhibit both internal and external coherence that grants them value as indicators of clinical plausibility. Internally, they reveal a consistent pattern in which biological deficiencies and family/school burdens are associated with greater difficulties in key psychological domains. Externally, they align with previous scientific literature documenting links between nutrition and language, parental stress and adaptive behavior, as well as environmental exposures and sensory modulation. Nonetheless, the pilot nature of the study and the non-diagnostic hair methodology underscore that causality cannot be inferred. These findings should be interpreted as signals guiding more robust hypotheses to be tested in future multicenter trials with quantitative biomarkers and larger samples, within the framework of the NeuroEpigenCare project.

## 6. Scientific Discussion and Limitations

### 6.1 Cautious Interpretation

The results of this pilot study support the feasibility and applicability of the BPSConnect protocol in a real clinical context, both in terms of family acceptability and the coherence of the findings with prior scientific literature. Integrating three dimensions traditionally approached independently biological, psychological, and social makes it possible to construct a more comprehensive explanatory framework for the experiences of children with ASD and the factors influencing their development. The identification of plausible associations between nutritional deficiencies, particularly vitamin D and iron, and socio-communicative and attentional difficulties, as well as the link between electromagnetic field (EMF) exposure and sensory alterations, provides evidence suggesting that biological and environmental factors may modulate the expression of ASD core symptoms. Similarly, the associations found between parental stress, school conditions, and adaptive functioning confirm that the child’s development cannot be analyzed in isolation from their family and educational contexts.

A noteworthy aspect of this study is its innovative character. To the best of our knowledge, this pilot constitutes the first European experience integrating exploratory epigenetic biomarkers, standardized psychometric scales, and social factors within a single screening protocol. This three-dimensional approach not only enhances the interpretation of ASD symptoms from an integrative perspective but also addresses the fragmentation typically observed in research, where biological, psychological, and social domains are often examined separately. By situating correlations within a unified framework, BPSConnect offers the potential to generate more holistic and clinically applicable evidence, providing professionals and families with a clearer understanding of the multifactorial origins of symptoms.

The relevance of this innovation is reinforced when comparing the preliminary findings with those of previously published international multicenter studies, which have demonstrated the influence of nutritional deficiencies, environmental factors, and parental burden on the developmental outcomes of children with ASD across different cultural contexts (Bener et al., 2017; Hayes & Watson, 2013; Gialloreti et al., 2022). The consistency between the signals detected in this pilot and the results reported in such studies lends credibility to the methodological direction adopted and suggests that BPSConnect could become an exportable and comparable model for future large-scale multicenter research.

Nonetheless, these findings must be interpreted with caution. The pilot nature of the study implies that the observed associations should not be taken as conclusive evidence of causality, but rather as preliminary signals guiding the formulation of more robust hypotheses. The cross-sectional design limits the ability to establish temporal relationships, and the moderate magnitude of the correlations although consistent with the literature requires validation through larger samples and more rigorous methodologies. Even so, the primary value of this pilot lies in demonstrating that it is feasible to operationalize a bio-psycho-social model within an applicable clinical protocol, representing a significant step forward in the comprehensive evaluation of ASD and laying the foundation for future multicenter developments with international impact.

### 6.2 Limitations

The methodological limitations of this study must be explicitly acknowledged to contextualize the findings and delineate the scope of the conclusions.

First, the small sample size (n=25) restricts the statistical power of the analyses, prevents the application of more complex multivariate adjustments, and limits the generalizability of the findings to broader and more diverse populations.

Second, the use of capillary biomarkers obtained through the Cell Wellbeing platform must be considered exploratory, as this is a non-quantitative method not validated for diagnostic purposes. While its potential as a preventive screening technique is promising due to its non-invasive nature and family acceptability, results require confirmation through gold-standard reference methods.

Third, the statistical models applied were constrained by the sample size, which limited the possibility of employing more sophisticated adjusted models to control for relevant confounders such as age, sex, baseline ASD severity, or medical comorbidities. In addition, the absence of a control group prevented comparison with children without ASD or with other neurodevelopmental disorders, which would have allowed for the assessment of the model’s specificity. Finally, the cross-sectional nature of the study restricts the capacity to establish longitudinal trajectories of change, preventing insight into how associations across dimensions evolve over time or under different interventions.

*This manuscript is a preprint and has not been peer-reviewed. Results are preliminary and should not guide clinical practice*.

### 6.3 Clinical and Social Relevance

Despite these limitations, the clinical and social relevance of the findings is evident.

First, social variables particularly parental stress and educational conditions emerged as critical modulators of adaptive functioning, behavioral difficulties, and school performance. This finding underscores the need for ASD approaches to transcend the individual level and adopt a systemic model integrating interventions targeting both families and schools. The fact that such a high proportion of caregivers reported elevated levels of stress and anxiety highlights the necessity of designing psychosocial support programs aimed at reducing family burden, fostering resilience, and indirectly improving the child’s developmental outcomes.

Second, the biological findings reinforce the importance of considering nutrition and metabolism as relevant factors in neurodevelopment. The identification of vitamin D and iron deficiencies in a substantial proportion of participants, and their associations with key domains such as socio-communication, language, and executive functions, aligns with prior studies linking micronutrient deficiencies to poorer ASD prognosis (Bener et al., 2017; Frye et al., 2020). These results suggest that nutritional assessment should become part of routine clinical practice in ASD management, not as a substitute for behavioral and educational interventions but as a necessary complement to optimize outcomes.

Finally, the environmental dimension represented in this study by EMF exposure although still underexplored in the literature, opens an emerging line of research that may shed light on individual variability in sensory sensitivity. While preliminary, this finding highlights the need for future studies to incorporate more rigorous measurements of environmental exposures and to examine their possible interactions with biological predispositions and family factors.

### 6.4 Need for Larger Studies (NeuroEpigenCare)

Although consistent with existing literature, the preliminary findings of this pilot must be interpreted with caution due to the exploratory nature of the design, the limited sample size, and the use of non-quantitative capillary biomarkers. Confirming these results requires larger-scale studies capable of overcoming current limitations and moving toward a robust validation of the BPSConnect model. In this context, the NeuroEpigenCare project is conceived as a necessary and strategic next phase, aimed at consolidating a comprehensive screening protocol with full clinical and scientific validity.

First, multicenter validation is essential to ensure the generalizability of results across different clinical, geographic, and socio-educational contexts. Involving multiple reference centers will allow for the recruitment of larger and more heterogeneous samples, control for variability linked to cultural and environmental factors, and assess the model’s applicability in diverse populations. This approach will also facilitate inter-center comparisons and the standardization of procedures conditions required for considering future transfer of the model into international clinical practice.

A second critical aspect is the incorporation of quantitative biomarkers obtained through standardized techniques with recognized diagnostic validity. In contrast to the exploratory capillary analysis used in this pilot, future studies must include serum determinations of vitamin D (25-OH-D), iron, and trace elements via reference methods such as LC-MS/MS or ICP-MS, thereby ensuring accuracy and reliability of biological data. Likewise, environmental exposure should be evaluated through objective dosimetric measures of electromagnetic fields and biomonitoring of heavy metals, enabling more robust correlations between biological and environmental factors and psychological and social profiles.

Similarly, the psychometric properties of the BPSConnect checklists must undergo systematic validation. Internal consistency of items (Cronbach’s α), temporal stability of responses (intraclass correlation coefficients, ICC), and convergent and divergent validity against standardized reference scales will need to be assessed. This process will not only guarantee the robustness of the instruments but also allow refinement and optimization for their digital implementation in clinical and educational settings.

Sample size calculation represents another fundamental requirement for confirmatory study design. The pilot provides initial estimates of variance and effect sizes useful for determining the number of participants needed to achieve adequate statistical power and minimize type II error risk. Unlike the exploratory character of this study, future research must be designed with confirmatory hypotheses and prespecified criteria, enabling the derivation of solid and generalizable conclusions.

Finally, longitudinal follow-up will be key to evaluating the stability and evolution of observed associations. Only through prospective designs will it be possible to determine whether biological, psychological, and social alterations identified at early stages predict developmental trajectories, therapeutic responses, and functional outcomes in children with ASD. Moreover, follow-up will allow assessment of the BPSConnect model’s sensitivity to detect changes after specific interventions, further consolidating its utility as a clinical evaluation and monitoring tool.

Taken together, the transition toward a multicenter study with quantitative biomarkers, rigorous psychometric validation, adequate sample size, and longitudinal design represents not only a methodological requirement but also an opportunity to position the BPSConnect model as an international reference in comprehensive ASD assessment. The NeuroEpigenCare project, conceived as the confirmatory phase of this pilot, therefore represents a decisive step toward establishing a reliable, scalable clinical instrument capable of guiding personalized interventions that integrate the biological, psychological, and social dimensions of neurodevelopment.

## 7. Conclusions

The analysis of the results obtained in this pilot study supports the feasibility of applying the BPSConnect protocol in real clinical settings and confirms that its methodological structure adequately addresses the needs of both professionals and participating families. Feasibility was demonstrated at several levels: first, the ability to complete assessments within clinically reasonable time frames; second, the capacity of the clinical team to apply the various tools without unnecessary duplication of processes; and third, the acceptance expressed by families and educational centers toward a procedure which, despite its complexity, was perceived as orderly, structured, and comprehensible. These elements are particularly relevant given that many screening or diagnostic tools ultimately fail not because of insufficient scientific rigor but due to practical barriers in implementation. In this case, the high completion rate and absence of incidents during administration support the conclusion that BPSConnect has a degree of viability that makes it a strong candidate for expansion in subsequent phases of clinical validation.

The preliminary clinical value of the findings should also be emphasized, as the protocol does not merely collect isolated information from different sources but integrates, within a single framework, three dimensions traditionally assessed separately: biological, psychological, and social. The ability to correlate biological signals such as vitamin D or iron deficiency with behavioral, linguistic, and attentional variables broadens the horizon of clinical interpretation, providing not only presence or absence of symptoms but also interaction patterns that can guide decision-making with greater precision. Likewise, the inclusion of questionnaires on family climate and school adaptation facilitates early identification of contexts of vulnerability that may condition the effectiveness of interventions. This integration is not trivial, as it breaks with the traditional tendency to compartmentalize the child’s experience into independent domains and instead promotes a systemic understanding of development.

The associations observed in this pilot are particularly revealing and plausible in light of the international scientific literature. The identification of links between vitamin D deficiency and socio-communicative difficulties, between low iron levels and problems in language and attention, and between parental stress and limitations in adaptive functioning aligns with previous studies that have described similar trajectories. These convergences with research conducted in other countries, using different methodologies, reinforce the model’s convergent validity and suggest that BPSConnect does not generate arbitrary findings but rather replicates patterns already recognized in the scientific literature. The congruence between clinical practice observations and previously published evidence is an indicator of robustness that strengthens confidence in the tool’s potential as a comprehensive screening model.

The social dimension emerges powerfully as an active modulator of ASD clinical expression. The finding that most caregivers experience high levels of stress, anxiety, and depression and that these conditions are associated with greater behavioral severity and poorer adaptive functioning in children demonstrates that child well-being cannot be dissociated from family well-being. Moreover, the lack of adequate educational supports, whether in terms of human resources, curricular adaptations, or inclusive methodologies, compounds family burden and contributes to intensifying the child’s observable symptomatology. Beyond confirming what has been previously described in the literature, these findings underscore the urgency of designing policies and intervention strategies that go beyond the clinical sphere and meaningfully involve educational and social systems. From this perspective, autism ceases to be a purely individual phenomenon and is reframed as a relational reality involving the family, the school, and the broader community.

Despite the relevance of these results, interpretation must remain cautious, acknowledging the limitations inherent to a pilot study. The sample size, restricted to 25 participants, precludes generalizable inferences, and the use of capillary epigenetic biomarkers although innovative must be regarded as exploratory, given the lack of diagnostic standardization and the need for validation with internationally recognized quantitative methodologies. Accordingly, the correlations observed should be understood as signals that justify future research but not as conclusive evidence of causality. Scientific rigor requires emphasizing that the results of this pilot do not establish deterministic relationships between biological deficits and clinical manifestations, but rather indicate associations consistent with trends described in larger studies.

The innovative character of this research lies not only in the simultaneous integration of biological, psychological, and social dimensions within a single instrument but also in the technological projection offered by its future implementation as a digital SaaS platform. Developing BPSConnect as software accessible to health professionals, educational specialists, and families represents a strategic advance toward the democratization of comprehensive diagnostic tools. Such a technological solution would enable automated data collection and analysis, generate clear and comprehensible reports, and provide parents and teachers with an integrated view of the multifactorial origins of each child’s symptoms. Digitalization would further facilitate longitudinal comparison of results, personalized follow-up, and the creation of population databases that could contribute to large-scale research and inform public policies in the field of neurodevelopment.

The most significant contribution of this pilot perhaps lies in its ability to lay the groundwork for the forthcoming NeuroEpigenCare project, which aims to consolidate protocol validity through a multicenter design, a larger sample, and the use of standardized quantitative biomarkers. The information generated in this preliminary phase provides indispensable operational parameters for designing that confirmatory trial, including estimates of administration time, identification of the most sensitive clinical domains, expected effect sizes in cross-dimensional correlations, and the definition of endpoints to guide subsequent analyses. In this sense, the pilot not only fulfills its exploratory purpose but also provides a clear and detailed roadmap for future research, facilitating resource planning, objective setting, and the structuring of robust methodologies.

This pilot should not be regarded as an endpoint but as a solid starting point that provides preliminary evidence of the relevance and potential of the BPSConnect model. The study shows that it is feasible to apply a comprehensive screening protocol in real clinical practice, that families and educators accept it positively, that the associations identified are consistent with the literature, and that the social dimension must occupy a central place in any approach to ASD. At the same time, it makes clear that methodological caution is indispensable and that the next steps must focus on validating the findings with more rigorous methodologies and larger, more diverse populations. The challenge posed by NeuroEpigenCare will be to transform these early signals into solid evidence capable of guiding clinical, educational, and social policy.

Within this framework, the development of a SaaS digital tool based on BPSConnect represents a high-impact innovation that may change the way autism is understood and addressed, providing professionals and families with unprecedented access to comprehensive assessments and clear reports illuminating the complex interplay between biology, psychology, and social context in the lives of children with ASD.

The BPSConnect model, validated in this pilot, thus emerges as a pioneering tool for future European and intercontinental multicenter studies in the field of neurodevelopment.

## Data Availability

Anonymized data supporting the findings of this study may be made available upon reasonable request to the corresponding author and under confidentiality agreement, given the sensitive nature of pediatric information and the intellectual protection of the instruments.

## Ethical Approval and Consent

Ethical approval: Research Ethics Committee of e-TherapyKids (Ref. BPS-TEA/2024-2025-11). Written informed consent was obtained from parents/legal guardians for all participants.

## Acknowledgments

The authors wish to thank the participating families for their time and commitment; the clinical and educational team for their operational support; and the individuals and entities who contributed to the fundraising campaign.

